# Seroprevalence of SARS-CoV-2-Specific IgG Antibodies Among Adults Living in Connecticut: Post-Infection Prevalence (PIP) Study

**DOI:** 10.1101/2020.08.04.20168203

**Authors:** Shiwani Mahajan, Rajesh Srinivasan, Carrie A. Redlich, Sara K. Huston, Kelly M. Anastasio, Lisa Cashman, Dorothy S. Massey, Andrew Dugan, Dan Witters, Jenny Marlar, Shu-Xia Li, Zhenqiu Lin, Domonique Hodge, Manas Chattopadhyay, Mark D. Adams, Charles Lee, Lokinendi V. Rao, Chris Stewart, Karthik Kuppusamy, Albert I. Ko, Harlan M. Krumholz

## Abstract

**Background:** A seroprevalence study can estimate the percentage of people with SARS-CoV-2 antibodies in the general population, however, most existing reports have used a convenience sample, which may bias their estimates.

**Methods:** We sought a representative sample of Connecticut residents, aged ≥18 years and residing in non-congregate settings, who completed a survey between June 4 and June 23, 2020 and underwent serology testing for SARS-CoV-2-specific IgG antibodies between June 10 and July 29, 2020. We also oversampled non-Hispanic Black and Hispanic subpopulations. We estimated the seroprevalence of SARS-CoV-2-specific IgG antibodies and the prevalence of symptomatic illness and self-reported adherence to risk mitigation behaviors among this population.

**Results:** Of the 567 respondents (mean age 50 [±17] years; 53% women; 75% non-Hispanic White individuals) included at the state-level, 23 respondents tested positive for SARS-CoV-2-specific antibodies, resulting in weighted seroprevalence of 4.0 (90% confidence interval [CI] 2.0–6.0). The weighted seroprevalence for the oversampled non-Hispanic Black and Hispanic populations was 6.4% (90% CI 0.9–11.9) and 19.9% (90% CI 13.2–26.6), respectively. The majority of respondents at the state-level reported following risk mitigation behaviors: 73% avoided public places, 75% avoided gatherings of families or friends, and 97% wore a facemask, at least part of the time.

**Conclusions:** These estimates indicate that the vast majority of people in Connecticut lack antibodies against SARS-CoV-2 and there is variation by race/ethnicity. There is a need for continued adherence to risk mitigation behaviors among Connecticut residents to prevent resurgence of COVID-19 in this region.

## INTRODUCTION

Connecticut was one of the first states in the United States (US) to be severely affected by Coronavirus Disease 2019 (COVID-19), with its first confirmed case of COVID-19 in early March. While almost 43,000 cases and 4,000 deaths were reported by June,^1^ a seroprevalence study, which estimates the percentage of people with SARS-CoV-2 antibodies, may provide a more accurate estimate of the percent of Connecticut population with evidence of a prior infection from COVID-19.

Prior seroprevalence studies have estimated the spread of COVID-19 in the US.^2-8^ However, the majority have taken advantage of blood samples collected for other reasons or used a convenience sample, which limits their representativeness. The Centers for Disease Control and Prevention (CDC) conducted a seroprevalence survey in Connecticut using blood specimens collected at commercial laboratories.^8^ However, these specimens were produced as part of routine or sick visit, representing a biased sample. Moreover, this effort did not provide the reason for the blood collection nor information about recent symptomatic illness, underlying conditions, or relevant risk-mitigation behaviors, which may help predict detection of antibodies against SARS-CoV-2.

Accordingly, with support from the Connecticut Department of Public Health (DPH) and the CDC, we conducted the Post-Infection Prevalence (PIP) Study, a public health surveillance project to determine the seroprevalence of SARS-CoV-2 among adults residing in community-non-congregate settings in Connecticut before June. Specifically, we sought to understand prior spread at the state-level; collect information about symptomatic illness, risk factors for virus infection, and self-reported adherence to risk mitigation behaviors; compare our seroprevalence estimates to available Connecticut estimates; and provide targeted estimates for the non-Hispanic Black and Hispanic populations.

## METHODS

### Study cohort

For the state-level seroprevalence estimate, from June 4 to June 23, 2020 we enrolled 735 adults residing in non-congregate settings (i.e. excluding individuals living in long-term care facilities, assisted living facilities, nursing homes, and prisons or jails) in Connecticut, aged ≥18 years, using a dual-frame Random Digit Dial (RDD) methodology.^9^ Additionally, from June 23 to July 22, 2020 we oversampled non-Hispanic Black (n=269) and Hispanic (n=341) individuals to provide more accurate estimates for these subpopulations. Details of the sample size calculation and RDD methodology are described in **eMethods 1**. Details of participant recruitment are described in **eMethods 2**. We contacted a total of 7305 respondents at the state-level, and successfully completed 735 interviews. We contacted a total of 12,508 respondents for the oversampled subpopulations, of whom 457 completed interviews.

The study was deemed not to be research by the Institutional Review Board at Yale University because of the public health surveillance activity exclusion and was approved by the Institutional Review Board at Gallup.

### Survey components

Individuals selected were provided study details, and informed consent was obtained from all participants by trained interviewers. Participants were interviewed using a questionnaire that collected information on demographics, social determinants of health, history of influenza-like-illness, symptoms experienced, and other COVID-19-related topics. The average survey time was 15 minutes.

### Specimen collection and serology testing

Within 24-48 hours of completing the interview, respondents were contacted to schedule their blood draw appointment at their nearest Quest Diagnostics Patient Service Center (PSC). Up to 5 attempts were made to each household where the participant agreed to be tested. Upon confirmation that the participant had completed the test, an incentive payment of $50 was sent as a gift card via email or mail. Beginning July 17, 2020, we offered participants an additional $50 (for a total compensation of $100) to incentivize completion of the serology test.

Of the 735 participants enrolled in the state-level estimate, 25 participants refused to participate when re-contacted for scheduling and 567 participants completed serology testing at 93 Quest Diagnostics PSCs throughout Connecticut between June 10 and July 29, 2020 (**eFigure 1**). Of the total 341 Hispanic and 269 non-Hispanic Black participants enrolled for the oversample estimate, 171 and 148 participants, respectively, completed serology testing (**eFigure 2**). The distribution of the timing of the blood draws is shown in **eFigure 3**.

Sera was obtained from samples collected in BD Hemogard serum separator tubes. All samples were processed at the Quest Diagnostics Marlborough Laboratory. Samples were run at room temperature using the primary collection tube. We measured IgG SARS-CoV-2 antibodies using Ortho-Clinical Diagnostics Vitros anti-SARS-CoV-2 IgG test, which detects antibodies against the spike glycoprotein of the virus.^10^ Antibody levels were expressed as the ratio of the chemiluminescence signal over the cutoff value, with a value ≥1.00 reported as positive.^11^ The Ortho Vitros IgG test had a reported sensitivity and specificity of 90% and 100%, respectively.^10^

We validated the sensitivity of this test in a small subset of SARS-CoV-2 positive patients (n=36) with variable disease severity, using reverse transcription polymerase chain reaction testing as the gold standard.^12^

Additionally, given the concern about the accuracy of serology tests,^13^ we re-tested the negative samples from 5 high risk cities of Connecticut (i.e. Bridgeport, Hartford, New Haven, Stamford, and Waterbury) with the Abbott Architect SARS-CoV-2 IgG test that detects antibodies aimed at a different SARS-CoV-2 antigen (nucleocapsid protein).^14^

Finally, Quest Diagnostics provided results for all SARS-CoV-2 serology tests conducted throughout Connecticut in the same time period (i.e. June 10 and July 29, 2020) for comparison.

### Statistical analysis

The sample data were weighted to approximate the Connecticut population (details described in **eMethods 3)**. Briefly, the base weight assigned to each completed survey was derived as the product of inverse of the probability of selection and non-response adjustment. Next, post-stratification weighting adjustments were made to account for residual non-response and to match the weighted sample estimates to known population characteristics for Connecticut. Post-stratification weighting for state-level sample was carried out using raking (or Iterative Proportional Fitting) procedures to adjust for age, gender, race/ethnicity, and education. The categories chosen for weighting the oversample subpopulations were different from what was used for the state-level adjustments due to lower available sample sizes. To reduce the effect of extreme weights on sampling variance, final weights were trimmed. The margin of error (MOE) for this study was calculated at the 90% confidence level (CI) taking into consideration the design effect introduced by variability of weights on each survey estimate. Overall study design effect as estimated by the Kish approximation equals 1.83, however, it varies by each survey estimate.

Next, the unweighted seroprevalence was calculated for both the overall state-level sample and the oversampled non-Hispanic Black and Hispanic subgroups. Finally, we estimated the weighted state-level seroprevalence and the MOE of these estimates, both overall and for subgroups with sufficient sample size. Subgroups with sample sizes <30 were too small to calculate accurate estimates and were thus not reported. We also estimated the MOE at 95% CI for the state-level estimates as a secondary outcome. We reported the weighted seroprevalence for non-Hispanic Black and Hispanic subgroups separately.

All statistical analyses were performed using SPSS 24.0 (SPSS, Inc. Chicago, IL) and R version 4.0.2. We considered 2-sided P-values <0.05 as statistically significant.

## RESULTS

### Population characteristics for the state-level sample

The final state-level sample included 567 respondents who completed both the survey and the serology test. The mean age of the weighted sample was 50.1 (±17.2) years, 53% were women, and the majority (75%) were non-Hispanic White individuals. Other weighted and unweighted characteristics of the study sample are reported in Table 1.

**Table 1.** Sociodemographic and clinical characteristics of adults included in the study for the state-level estimate.

| **Characteristics** | **Unweighted**<br>**N** | **Unweighted**<br>**Proportion,**<br>**%** | **Weighted**<br>**Proportion,**<br>**%** | **Target**<br>**Percentage<sup>1</sup>,**<br>**%** |
| --- | --- | --- | --- | --- |
| **Overall** | 567 | - | 567 | - |
| **Age group, years** |  |  |  |  |
| 18-29 | 41 | 7.2% | 13.1% | 19.9% |
| 30-44 | 90 | 15.9% | 26.7% | 22.9% |
| 45-54 | 113 | 19.9% | 18.6% | 17.5% |
| 55-64 | 134 | 23.6% | 18.6% | 18.1% |
| ≥65 | 187 | 33.0% | 23.0% | 21.6% |
| **Sex** |  |  |  |  |
| Men | 244 | 43.0% | 46.6% | 48.1% |
| Women | 323 | 57.0% | 53.4% | 51.9% |
| **Race/Ethnicity** |  |  |  |  |
| Hispanic | 49 | 8.6% | 13.0% | 14.4% |
| Non-Hispanic White | 470 | 82.9% | 74.9% | 69.4% |
| Non-Hispanic Black | 37 | 6.5% | 9.6% | 9.8% |
| Non-Hispanic Asian | 9 | 1.6% | 1.2% | 4.7% |
| Non-Hispanic Other | 5 | 0.9% | 1.7% | 1.7% |
| **Education level** |  |  |  |  |
| Less than high school | 7 | 1.2% | 3.7% | 9.3% |
| High school or GED | 79 | 13.9% | 33.2% | 27.4% |
| Some college | 131 | 23.1% | 23.9% | 26.5% |
| Bachelor's degree or more | 350 | 61.7% | 39.2% | 36.8% |
| **Income level** |  |  |  |  |
| Less than \$24,000 | 40 | 7.1% | 11.3% | N/A |
| \$24,000 to \$59,999 | 104 | 18.3% | 25.0% | N/A |
| \$60,000 to \$119,999 | 178 | 31.4% | 30.1% | N/A |
| \$120,000 or more | 195 | 34.4% | 26.8% | N/A |
| Don't know/Refused | 50 | 8.8% | 6.8% | N/A |
| **Health insurance** |  |  |  |  |
| Yes | 554 | 97.7% | 95.3% | 94.0% |
| No | 13 | 2.3% | 4.7% | 6.0% |
| **Employment status** |  |  |  |  |
| Employed full-time | 263 | 46.4% | 45.2% | 63.8% |
| Employed part-time | 56 | 9.9% | 10.0% |  |
| Unemployed | 43 | 7.6% | 10.6% | 3.5% |
| Retired/Student/Homemaker | 175 | 30.9% | 25.5% | N/A |
| Disabled | 0 | 0.0% | 0.0% | N/A |
| Unknown | 30 | 5.3% | 8.6% | N/A |
| **Essential job (exempt from**<br>**stay-at-home orders)** |  |  |  |  |
| Yes | 140 | 24.7% | 27.5% | N/A |
| No | 169 | 29.8% | 24.8% | N/A |
| Don't know/Refused/Not employed | 258 | 45.5% | 47.7% | N/A |
| **Region/County** |  |  |  |  |
| Fairfield | 126 | 22.2% | 25.2% | 25.8% |
| Hartford | 157 | 27.7% | 24.1% | 24.9% |
| Litchfield | 42 | 7.4% | 5.5% | 5.2% |
| Middlesex | 34 | 6.0% | 5.0% | 4.7% |
| New Haven | 131 | 23.1% | 24.3% | 24.1% |
| New London | 41 | 7.2% | 7.8% | 7.6% |
| Tolland | 20 | 3.5% | 4.5% | 4.4% |
| Windham | 16 | 2.8% | 3.6% | 3.3% |
| **Type of home** |  |  |  |  |
| Mobile home | 2 | 0.4% | 1.0% | N/A |
| Single family house or townhouse | 447 | 78.8% | 69.7% | N/A |
| Apartment or condo | 112 | 19.8% | 28.1% | N/A |
| Group facility | 2 | 0.4% | 0.3% | N/A |
| Don't know/Refused | 4 | 0.7% | 0.9% | N/A |
| **Self-reported health status** |  |  |  |  |
| Excellent | 177 | 31.2% | 29.7% | N/A |
| Very good | 223 | 39.3% | 32.0% | N/A |
| Good | 128 | 22.6% | 26.9% | N/A |
| Fair | 33 | 5.8% | 9.2% | N/A |
| Poor | 6 | 1.1% | 2.3% | N/A |
| **Chronic conditions** |  |  |  |  |
| Diabetes | 64 | 11.3% | 12.2% | N/A |
| Asthma, COPD or another lung disease | 63 | 11.1% | 16.7% | N/A |
| Heart disease | 37 | 6.5% | 6.9% | N/A |
| Cancer | 72 | 12.7% | 10.7% | N/A |
| High blood pressure | 171 | 30.2% | 30.5% | N/A |
| Immune compromised | 46 | 8.1% | 8.5% | N/A |
| **Lived in Connecticut in past 12 weeks** |  |  |  |  |
| <6 weeks | 9 | 1.6% | 1.0% | N/A |
| 6-10 weeks | 13 | 2.3% | 1.9% | N/A |
| 11-12 weeks | 543 | 95.8% | 96.5% | N/A |
| Don't know/Refused | 2 | 0.4% | 0.6% | N/A |
| <sup>1</sup> Source for age, sex, race, ethnicity, education, employment, county targets: American Community Survey 2018. Source for health insurance: Reference information for health insurance coverage is obtained from the Current Population Survey estimates, 2018. Target percentage is based on expected proportions for a perfectly random sample, based on credible external sources.<br>Abbreviations: COPD, Chronic Obstructive Pulmonary Disease; GED, General Educational Development test; N/A, Not Available. |  |  |  |  |

Comparison of the unweighted demographic distribution of individuals who completed only the survey with those who completed both the survey and the antibody test has been provided in **eTable 1**. While the 2 groups were not significantly different in regional representation, a significantly higher number of younger, Hispanic and non-Hispanic Black individuals did not complete blood testing. However, our weighted study sample was closer to the target sample in the distribution of subgroups by age, sex, race/ethnicity, education level, and health insurance (**Table 1**).

### Symptoms and risk mitigation behaviors at the state-level

As shown in **Table 2**, cough, diarrhea, fever, sore throat and new onset loss of taste or smell was reported by 18%, 16%, 9%, 10%, and 5% respondents, respectively, at some point between March and June. About 16% individuals reported being tested for coronavirus previously, and of these, 12% reported testing positive.

**Table 2.** Prevalence of symptomatic illness, risk factors for possible exposure, and adherence to social-distancing behaviors since March 1, 2020 among the state-level population.

| Characteristics | Unweighted<br>N | Unweighted<br>Proportion, % | Weighted Proportion,<br>% (MOE) |
| --- | --- | --- | --- |
| **Symptoms** |  |  |  |
| Fever | 43 | 7.6% | 8.7% ($\pm 2.7$) |
| Cough | 84 | 14.8% | 18.3% ($\pm 3.5$) |
| Sore throat | 54 | 9.5% | 10.1% ($\pm 2.7$) |
| New loss of taste or smell | 25 | 4.4% | 4.8 ($\pm 2.0$) |
| Diarrhea | 70 | 12.3% | 15.9% ($\pm 3.2$) |
| **Risk Factors/Behaviors** |  |  |  |
| Received coronavirus test | 90 | 15.9% | 15.7% ($\pm 3.5$) |
| Tested positive for coronavirus | 11 | 1.9% | 1.9% ($\pm 1.4$) |
| Anyone in household (other than respondent) had symptoms of coronavirus | 54 | 9.5% | 10.2% ($\pm 2.7$) |
| Anyone in household (other than respondent) tested positive for coronavirus | 16 | 2.8% | 3.7% ($\pm 1.8$) |
| Avoided going to public places, such as stores or restaurants | 422 | 74.4% | 72.8% ($\pm 4.1$) |
| Avoided small gatherings of people, with family or friends | 426 | 75.1% | 75.3% ($\pm 4.0$) |
| Worked from home (among all respondents, regardless of employment status) | 223 | 39.3% | 31.4% ($\pm 4.1$) |
| Worn a mask on your face when outside your home | 557 | 98.2% | 96.9% ($\pm 1.6$) |
| Traveled by airplane | 39 | 6.9% | 6.5% ($\pm 2.6$) |
| Traveled using public transportation, such as bus or train | 19 | 3.4% | 5.2% ($\pm 2.0$) |
| Abbreviations: MOE, Margin of Error at the 90% confidence level |  |  |  |

The majority of respondents reported following risk mitigation practices, at least some of the time, since March, with 73% reporting having avoided public places and 75% reporting having avoided gatherings of family and friends. Notably, 97% respondents reported wearing mask outside their home at least part of the time. About 31% of all respondents reported having worked from home at least part of the time, representing 57% of working respondents. We compared the prevalence of symptomatic illness and risk mitigation behaviors among individuals who completed only the survey with those who completed the survey and the antibody test in **eTable 2**.

### Seroprevalence of SARS-CoV-2 antibodies at the state-level

Seroprevalence estimates are shown in **Table 3**. Overall, 23 respondents tested positive for SARS-CoV-2 antibodies, yielding a weighted seroprevalence of 4.0% (90% CI 2.0–6.0). Among individuals who reported having symptomatic illness, those with fever, cough, sore throat, and diarrhea had a weighted seroprevalence of 32.4% (90% CI 15.1–49.7), 11.4% (90% CI 2.8–20.0), 10.3% (90% CI 0.0–21.0), and 6.9% (90% CI 0.0–14.4), respectively. Among the 25 individuals who reported loss of taste or smell, 14 individuals tested positive for SARS-CoV-2-specific antibodies.

**Table 3.**
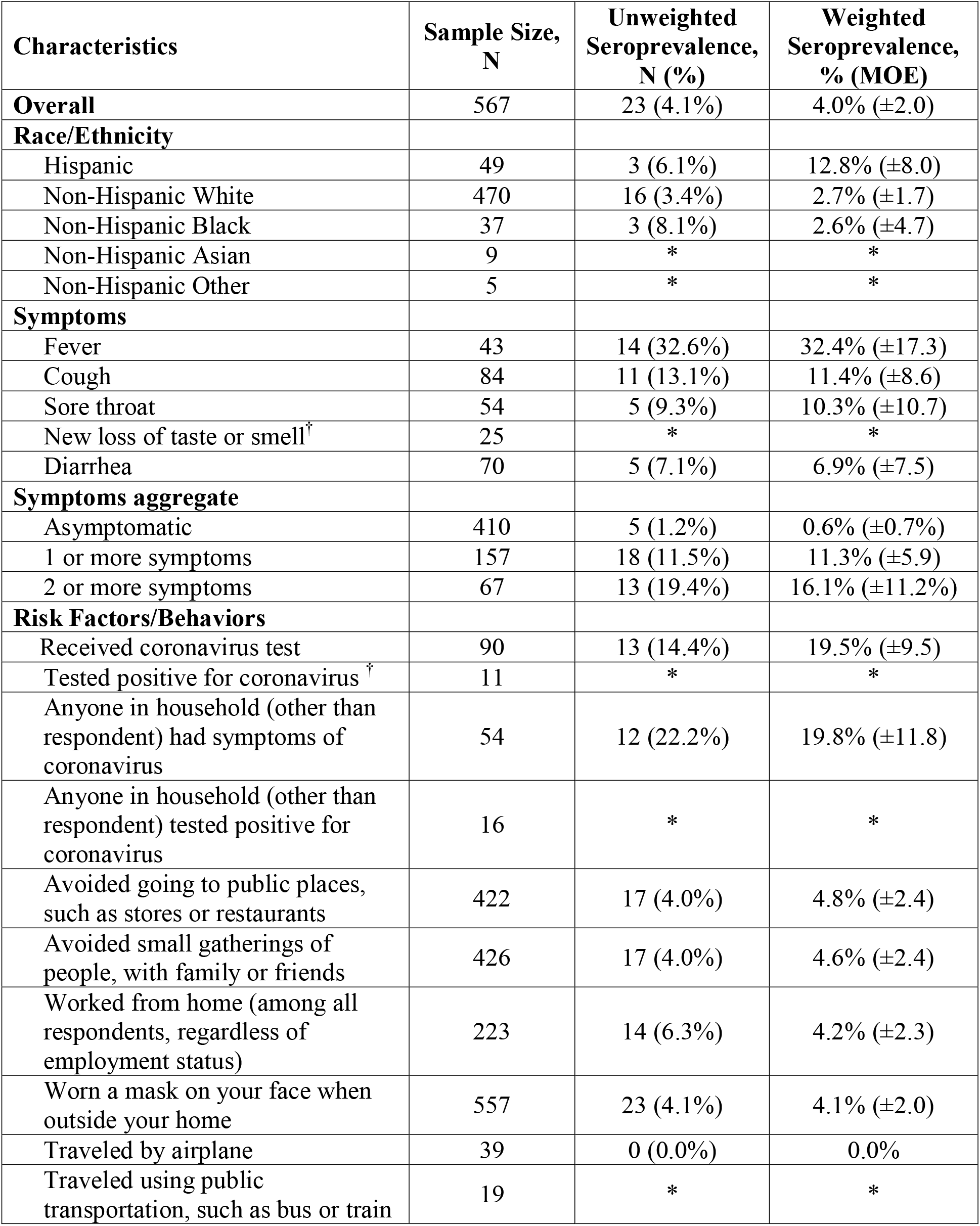
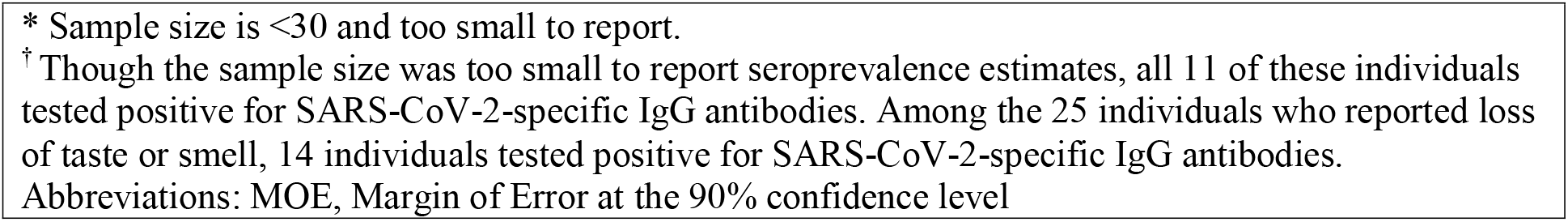
Unweighted and weighted state-level seroprevalence of SARS-CoV-2-specific IgG antibodies among adults in Connecticut, overall and by symptoms and risk factors and behaviors.

| Characteristics | Sample Size,<br>N | Unweighted<br>Seroprevalence,<br>N (%) | Weighted<br>Seroprevalence,<br>% (MOE) |
| --- | --- | --- | --- |
| **Overall** | 567 | 23 (4.1%) | 4.0% ($\pm 2.0$) |
| **Race/Ethnicity** |  |  |  |
| Hispanic | 49 | 3 (6.1%) | 12.8% ($\pm 8.0$) |
| Non-Hispanic White | 470 | 16 (3.4%) | 2.7% ($\pm 1.7$) |
| Non-Hispanic Black | 37 | 3 (8.1%) | 2.6% ($\pm 4.7$) |
| Non-Hispanic Asian | 9 | * | * |
| Non-Hispanic Other | 5 | * | * |
| **Symptoms** |  |  |  |
| Fever | 43 | 14 (32.6%) | 32.4% ($\pm 17.3$) |
| Cough | 84 | 11 (13.1%) | 11.4% ($\pm 8.6$) |
| Sore throat | 54 | 5 (9.3%) | 10.3% ($\pm 10.7$) |
| New loss of taste or smell<sup>†</sup> | 25 | * | * |
| Diarrhea | 70 | 5 (7.1%) | 6.9% ($\pm 7.5$) |
| **Symptoms aggregate** |  |  |  |
| Asymptomatic | 410 | 5 (1.2%) | 0.6% ($\pm 0.7$) |
| 1 or more symptoms | 157 | 18 (11.5%) | 11.3% ($\pm 5.9$) |
| 2 or more symptoms | 67 | 13 (19.4%) | 16.1% ($\pm 11.2$) |
| **Risk Factors/Behaviors** |  |  |  |
| Received coronavirus test | 90 | 13 (14.4%) | 19.5% ($\pm 9.5$) |
| Tested positive for coronavirus<sup>†</sup> | 11 | * | * |
| Anyone in household (other than respondent) had symptoms of coronavirus | 54 | 12 (22.2%) | 19.8% ($\pm 11.8$) |
| Anyone in household (other than respondent) tested positive for coronavirus | 16 | * | * |
| Avoided going to public places, such as stores or restaurants | 422 | 17 (4.0%) | 4.8% ($\pm 2.4$) |
| Avoided small gatherings of people, with family or friends | 426 | 17 (4.0%) | 4.6% ($\pm 2.4$) |
| Worked from home (among all respondents, regardless of employment status) | 223 | 14 (6.3%) | 4.2% ($\pm 2.3$) |
| Worn a mask on your face when outside your home | 557 | 23 (4.1%) | 4.1% ($\pm 2.0$) |
| Traveled by airplane | 39 | 0 (0.0%) | 0.0% |
| Traveled using public transportation, such as bus or train | 19 | * | * |

\* Sample size is <30 and too small to report.
† Though the sample size was too small to report seroprevalence estimates, all 11 of these individuals tested positive for SARS-CoV-2-specific IgG antibodies. Among the 25 individuals who reported loss of taste or smell, 14 individuals tested positive for SARS-CoV-2-specific IgG antibodies.
Abbreviations: MOE, Margin of Error at the 90% confidence level

Asymptomatic individuals had significantly lower weighted seroprevalence 0.6% (90% CI 0.0–1.3) compared with the overall state estimate, while those with >1 and >2 symptoms had a seroprevalence of 11.3% (90% CI 5.4–17.2) and 16.1% (90% CI 4.9–27.3), respectively (**Table 3**). The comparisons between other subgroups and the state estimates are presented in **eTable 3**. Additionally, seroprevalence estimates at 95% MOE have also been shown in **eTable 3**.

Among the 143 negative samples from 5 high risk cities of Connecticut that were re-tested with Abbott Architect serology assay, 142 (99.3%) samples tested negative. Additionally, of the total 25,274 antibody tests conducted by Quest Diagnostics in Connecticut during this time period, 2072 (8.4%) samples tested positive. Of the 11 respondents who reported testing positive for coronavirus, all tested positive for antibodies.

### Characteristics and seroprevalence estimates among non-Hispanic Black and Hispanic subpopulations

For the subpopulation estimate, the final sample included 171 Hispanic (39.9 [±15.5] years and 51% women) and 148 non-Hispanic Black (46.4 [±13.0] years and 56% women) adults (**eTable 4**). Fever, cough, sore throat, diarrhea, and new loss of taste or smell was reported by 11%, 17%, 15% 10%, and 8% of Hispanic participants and 4%, 10%, 5%, 4%, and 6% of Black participants (**Table 4**). About 37% of Hispanic and 31% of non-Hispanic Black individuals reported receiving a coronavirus test previously and nearly 6% of Hispanic and 4% non-Hispanic Black individuals reported testing positive for coronavirus. The prevalence of symptomatic illness and risk mitigation behaviors among individuals who completed only the survey has been compared with those who completed both the survey and the antibody test in **eTable 5**.

**Table 4.** Prevalence of symptomatic illness, risk factors for possible exposure, and adherence to social-distancing behaviors since March 1, 2020 among non-Hispanic Black and Hispanic subpopulation.

| Characteristics | Hispanic subpopulation |  |  | Non-Hispanic Black subpopulation |  |  |
| --- | --- | --- | --- | --- | --- | --- |
|  | Unweighted<br>N | Unweighted<br>Proportion,<br>% | Weighted<br>Proportion,<br>% (MOE) | Unweighted<br>N | Unweighted<br>Proportion,<br>% | Weighted<br>Proportion,<br>% (MOE) |
| **Overall** | 171 | N/A | N/A | 148 | N/A | N/A |
| **Symptoms** |  |  |  |  |  |  |
| Fever | 16 | 9.4% | 10.8% ($\pm 5.6$) | 7 | 4.7% | 3.9% ($\pm 3.6$) |
| Cough | 31 | 18.1% | 17.4% ($\pm 6.4$) | 18 | 12.2% | 10.1% ($\pm 6.4$) |
| Sore throat | 30 | 17.5% | 15.0% ($\pm 6.1$) | 8 | 5.4% | 4.7% ($\pm 4.6$) |
| New loss of taste or smell | 15 | 8.8% | 7.8% ($\pm 4.3$) | 8 | 5.4% | 4.2% ($\pm 3.8$) |
| Diarrhea | 25 | 14.6% | 10.2% ($\pm 5.5$) | 11 | 7.4% | 5.7% ($\pm 4.7$) |
| **Risk Factors/Behaviors** |  |  |  |  |  |  |
| Received coronavirus test | 64 | 37.4% | 36.9 ($\pm 9.6$) | 54 | 36.5% | 31.0% ($\pm 10.5$) |
| Tested positive for coronavirus<br>(out of all participants,<br>regardless of prior testing) | 10 | 5.8% | 6.2% ($\pm 3.8$) | 9 | 6.1% | 3.9% ($\pm 4.7$) |
| Anyone in household (other<br>than respondent) had symptoms<br>of coronavirus | 28 | 16.4% | 20.6% ($\pm 6.8$) | 6 | 4.1% | 2.1% ($\pm 2.1$) |
| Anyone in household (other<br>than respondent) tested positive<br>for coronavirus | 15 | 8.8% | 9.3% ($\pm 4.6$) | 3 | 2% | 2.0% ($\pm 2.7$) |
| Avoided going to public places,<br>such as stores or restaurants | 139 | 81.3% | 79.2% ($\pm 7.6$) | 96 | 64.9% | 63.8% ($\pm 10.5$) |
| Avoided small gatherings of<br>people, with family or friends | 140 | 81.9% | 81.6% ($\pm 6.9$) | 108 | 73% | 75.4% ($\pm 9.2$) |
| Worked from home (among all<br>respondents, regardless of<br>employment status) | 42 | 24.6% | 11.8% ($\pm 5.7$) | 48 | 32.4% | 17.8% ($\pm 9.0$) |
| Worn a mask on your face<br>when outside your home | 168 | 98.2% | 97.7 ($\pm 2.8$) | 145 | 98% | 96.5% ($\pm 4.0$) |
| Traveled by airplane | 11 | 6.4% | 4.8% ($\pm 3.3$) | 6 | 4.1% | 4.0% ($\pm 4.8$) |
| Traveled using public transportation, such as bus or train | 11 | 6.4% | 13.1% ($\pm 5.5$) | 16 | 10.8% | 23.7% ($\pm 7.5$) |
| Abbreviations: MOE, Margin of Error at the 90% confidence level |  |  |  |  |  |  |

The weighted seroprevalence among the Hispanic and non-Hispanic Black subpopulation, derived from both the random state sample and the oversample, was 19.9% (90% CI 13.2–26.6) and 6.4% (90% CI 0.9–11.9) respectively. The seroprevalence estimate for the Hispanic group was significantly higher than the overall state-level estimate.

## DISCUSSION

Our study primarily shows that despite Connecticut being an early COVID-19 hotspot, the vast majority of people in Connecticut lack detectable antibodies to SARS-CoV-2. In addition, individuals who reported having symptomatic illness between March and June of 2020 had higher seroprevalence rates, but over 90% of these individuals did not have SARS-CoV-2-specific IgG antibodies. Also, a high percentage of people interviewed reported following risk mitigation strategies, which may be partly responsible for the reduction in the number of new COVID-19 cases being reported in Connecticut. Finally, the Hispanic subpopulation had a higher prevalence of SARS-CoV-2-specific antibodies as compared with the overall state-level estimate, suggesting that the burden of disease was higher in this subgroup.

Our findings are consistent with other reports of more selected Connecticut populations. The CDC conducted a seroprevalence study using commercial laboratory data and reported a seroprevalence of 4.9% (95% CI 3.6–6.5) between April 26 and May 3 and 5.2% (95% CI 3.8–6.6) between May 21 and May 26 in Connecticut.^2,8^ However, these estimates were from people who had blood specimens tested for reasons unrelated to COVID-19, such as for a routine or sick visit, and as such would be expected to be biased higher than estimates for the general population. Similarly, data for all antibody tests conducted by Quest Diagnostics in Connecticut between June 10 and July 29, showed a seropositivity rate of 8.4%. Since these estimates were also among people who had a serology test done at a commercial laboratory, it is likely that these specimens were drawn from individuals who were more likely to suspect prior disease exposure than the general population.

Overall, our findings are consistent with other reports of population-level seroprevalence of SARS-CoV-2 in Europe and the US, although the burden of disease in these regions may have varied. A recent report from Spain,^15^ reported a seroprevalence of 4.6% (95% CI 4.3–5.0) and a population-based study from Switzerland,^16^ reported SARS-CoV-2 antibodies in <10% of the population. Reports from regions within the US have also shown similar numbers. A recent report from Indiana^5^ found a seropositivity rate of 1.01% (95% CI, 0.76–1.45) and a community seroprevalence survey from Atlanta^4^ estimated seroprevalence of 2.5% (95% CI, 1.4–4.5). Our findings of a higher burden of SARS-CoV-2 antibodies among Hispanic subgroups is also consistent with prior reports demonstrating that minority populations have been disproportionately affected by COVID-19.^5,17^

There are several explanations for why our state-level estimates are lower than what one might expect given that Connecticut had nearly 43,000 positive cases and 4,000 COVID-19 deaths by June 1, 2020. First, the majority of those deaths were among residents of congregate facilities. Second, the response and serology testing rates may have influenced the result. Only 7% of those contacted by phone completed the survey and blood test and the recruited population differed from the targets. However, this is a standard response rate in studies seeking representative populations and was considered in weighting the data. It is also possible that those who were more likely to have a positive test failed to complete the blood draw in higher proportions. However, this non-response was taken into account while weighting the sample. Third, there is some evidence suggesting a short-lived antibody response, especially among individuals with mild or asymptomatic illness,^18,19^ and it is possible that more people were infected who lost antibodies over time. However, recent studies suggest that the decline in this timeframe is small and antibody levels can remain stable for up to 120 days,^20,21^ and all 11 people who reported receiving a previous coronavirus test in our study tested positive for antibodies. Fourth, the accuracy of the serology tests has been a concern.^13^ However, 99% of the negative serology samples from the highest risk regions of Connecticut that we re-tested with Abbott Architect serology assay tested negative a second time.

Nevertheless, our findings are concordant with other studies in indicating that the vast majority of the population in Connecticut does not have detectable levels of antibodies against SARS-CoV-2. At present, we do not know whether anti-SARS-CoV-2 antibodies confer immunity. If such antibodies, as detected by ELISA, are a marker of immunity, then more than 95% of the people in Connecticut would be susceptible to the virus. Given low infection rates over the summer, these general estimates are still reasonable. As such, there is continued need for strong public health efforts encouraging Connecticut residents to adhere to risk mitigation behaviors so as to prevent a second wave of spread in the region.

### Conclusion

Our findings indicate that even in one of the early hotspots of the SARS-CoV-2 outbreak in the US, most of the population does not have detectable antibodies against SARS-CoV-2, and as such, remains vulnerable to infection. Also, there is notable variation by race/ethnicity. People likely need to continue to be vigilant about practices that can slow the spread in order to prevent resurgence of the virus in these regions.

## Data Availability

N/A

## ACKNOWLEDGEMENTS

We are thankful to Matt Cartter, Josh Geballe, and Deidre Gifford from the Connecticut Department of Public Health for their assistance with the funding. We are also thankful to Michael F. Murray, Saad B. Omer, Alan Gerber, Adam Wisnewski, Richard Torres, Nathan Grubaugh, Wade Schulz, Tesheia Johnson, Cesar Caraballo-Cordovez, Yuan Lu, Erica S. Spatz, and Karthik Murugiah from Yale University for their support. Finally, we are in debt to those who participated in the surveys and completed the serology test, and the many people at our organizations who spent countless hours ensuring the success of the project and contributing to the public’s health.

